# Neuroanatomical correlates of polygenic risk for Parkinson’s Disease

**DOI:** 10.1101/2022.01.17.22269262

**Authors:** Nooshin Abbasi, Christina Tremblay, Reza Rajimehr, Eric Yu, Ross D Markello, Golia Shafiei, Nina Khatibi, The ENIGMA-Parkinson’s study, Neda Jahanshad, Paul M. Thompson, Ziv Gan-Or, Bratislav Misic, Alain Dagher

**Affiliations:** Montreal Neurological Institute, McGill University, Montreal QC Canada; MRC Cognition and Brain Sciences Unit, University of Cambridge, Cambridge, UK; Department of Human Genetics, McGill University, Montreal QC Canada; Department of Biomedical Engineering, North Tehran Azad University, Tehran, Iran; Imaging Genetics Center, Mark and Mary Stevens Neuroimaging & Informatics Institute, Keck School of Medicine of the University of Southern California, Marina del Rey, California, USA

## Abstract

Parkinson ‘s Disease (PD) is heritable, however how genetic risk confers vulnerability remains mostly unknown. Here we use genetic and neuroimaging measures from 20,000 healthy adults from the UK Biobank to show that PD polygenic risk score (PRS) is associated with cortical thinning in a pattern that resembles cortical atrophy seen in PD. Conversely, PD PRS is associated with a global increase in cortical surface area. We also show that the genetically determined cortical thinning profile overlaps with the expression of genes associated with synaptic signaling, is dependent on anatomical connectivity and on regional expression of the most significant PD risk genes. Despite identical PRS distributions in males and females, only males show the associated brain features, possibly explaining the sex disparity in PD. We discuss potential mechanisms linking genetic risk to cortical thickness and surface area, and suggest that the divergent effects may reflect separate routes of genetic vulnerability.

## Introduction

Parkinson ‘s Disease (PD) is a progressive neurodegenerative movement disorder that usually presents after age 50. Age is the greatest risk factor, but there is a long prodromal phase prior to the onset of motor symptoms ^1^. The motor manifestations, bradykinesia, tremor, and rigidity are referred to as “parkinsonism” and are due to loss of dopamine neurons in the *substantia nigra pars compacta*. However, the pathological process affects the entire central nervous system ^2^, as evidenced by the presence of diffuse brain atrophy even in prodromal and *de novo* cases ^3,4^. Most cases of PD are thought to be due to misfolded pathogenic alpha-synuclein, whose accumulations are visible at post-mortem as Lewy bodies and neurites ^5,6^. Misfolded alpha-synuclein propagates through the brain via neuronal connections; its accumulation is associated with loss of certain neuronal populations (notably dopamine neurons), but also widespread loss of synapses with an associated diffuse pattern of brain atrophy on MRI ^4,7–10^.

Until recently, PD was thought to consist of either rare familial forms or cases of environmental origin. However, there is now clear evidence of genetic influence even in sporadic cases. Genome-wide association studies (GWAS) have identified numerous genetic variants with small cumulative effects, which can be aggregated into a polygenic risk score (PRS) that may explain up to 36% of PD heritability ^11^. Interestingly, many of the genes with rare mutations implicated in monogenic forms of PD also contribute to the polygenic risk. Many of these genes affect interrelated disease mechanisms such as protein homeostasis, autophagy and lysosomal function, and accumulation or clearance of abnormal alpha-synuclein isoforms ^12–14^. For example, three genes with significant signals in the GWAS are *SNCA*, which encodes alpha-synuclein, *LRRK2*, whose product is involved in autophagy and lysosomal function and influences alpha-synuclein mediated neurodegeneration ^15^, and *GBA*, which encodes a key lysosomal enzyme implicated in alpha-synuclein degradation ^13^. Mutations in *SNCA* and *LRRK2* are also causes of autosomal dominant PD, while *GBA* mutations are a risk-factor for PD ^16,17^. Note however that the function of most of the other genes contributing to the PRS remains unknown. The PD PRS also influences disease severity, as indicated by correlations with age at onset ^18^ and rate of cognitive and motor progression ^19^.

An important but unresolved question concerns how the potential risk genes translate into vulnerability to disease. PD likely results from an interaction of genetic, environmental, and stochastic causes. Genetic risk for PD may be associated with a vulnerability to the pathophysiological mechanisms that are associated with PD, namely alpha-synuclein accumulation and autophagy-lysosomal dysfunction. This could explain how a high genetic risk confers both vulnerability to PD and more severe disease. It may be possible to detect evidence of this vulnerability in the brains of healthy older individuals. Therefore, a first goal of the current work is to relate PD-PRS to brain morphometry in healthy adults.

Considerable evidence now supports a model of PD whereby misfolded alpha-synuclein propagates in a prion-like manner along neuronal connections ^20–22^. This is reflected in MRI studies in PD showing that neural connectivity shapes the pattern of brain atrophy ^4,23,24^. We therefore also sought to determine whether any regional brain patterns related to PD-PRS are explainable by brain connectivity.

We used genetic, neuroimaging, and behavioral data from the UK Biobank (UKB) ^25^ to uncover the neural and clinical correlates of a genetically determined high-risk state. We first show that the PD-PRS is associated with a pattern of cortical thickness variability that resembles the pattern of atrophy seen in PD patients. We then demonstrate that this pattern is explained by brain connectivity, supporting an underlying propagating process. Moreover, the PD-PRS brain pattern also overlaps spatially with the normal brain expression of the most impactful PD-risk related genes. Surprisingly, we also find that the PD-PRS is associated with greater cortical surface area, affecting the entire cortical mantle. These opposing effects on cortical thickness and surface area may suggest divergent mechanisms of genetic vulnerability to PD.

We also examined certain traits associated with PD to see if any neurobehavioral effects of genetic risk manifest in this population. Considerable evidence supports an association between PD and a reduced tendency for addictive behaviors including cigarette smoking and alcohol intake, sometimes occurring years before motor symptoms ^11,26^. This has been hypothesized to reflect reduced dopaminergic signaling. There may also be a positive association between PD and intelligence and educational attainment ^27^. For all these traits the direction and even existence of causal relationships is unclear ^28–30^. We therefore asked whether, in the entire UKB sample, there was a relation between the aforementioned traits and the PD-PRS. Finally, we were interested to see whether any PD-PRS effects could explain the known sex differences in the incidence and severity of PD ^31^.

## Results

### Genetic and Neuroimaging Data

We studied brain, genetic, and behavioral data of 29,101 healthy participants aged 45 to 82 years old (mean: 64.3) from the UKB ^25,32^. We applied the following exclusion criteria: history of bipolar or any neurological disorder including degenerative, vascular, traumatic, and infectious brain pathologies; first degree family history of Parkinson ‘s disease; body mass index (BMI) > 35; relation to another participant closer than cousin; genetic and self-reported sex mismatch; and non-European ancestry. We calculated PD-PRS by using the R package PRSice2 ^33^ with genetic risk data from the largest available Parkinson ‘s disease GWAS meta-analysis ^11^. For brain morphometry, we used MRI measures of cortical thickness and surface area from parcels of the Desikan-Killiany (DK) atlas ^34^ as described in ^35^. We also obtained a thorough list of potential imaging confounds including age, age-squared, sex, head motion in resting and task fMRI, date and date-squared for each UKB study site ^36^. Genotyping batch and the first fifteen genetic principal components, accounting for population ancestry stratification, were also added to the confound list ^37^.

### Association of PD-PRS with cortical thickness and surface area

We first performed a partial least squares (PLS) analysis ^38^, to assess whether a genetic composition of PD-PRS and population ancestry structure is able to explain any degree of variation in cortical thickness and surface area (Fig. 1). PLS is a multivariate approach based on singular value decomposition (SVD) of the data matrices to investigate the linear relationship between two sets of variables. To overcome the effects of the above-mentioned confounds, their contribution was regressed out from the brain maps prior to the PLS analysis. The significance of the covariance explained by each latent variable was obtained from non-parametric permutation testing, and bootstrapping was further employed to obtain a confidence interval for the contribution of each feature to the latent variables. Two significant latent variables were identified for both cortical thickness and surface area, in which the first latent variable was able to explain more than 10% (Fig. 1a) and 30% (Fig. 1f) of the variance in the two cortical measures, respectively. Bootstrap testing of the first latent variable highlighted a significant and major contribution of PD-PRS in determining the variation in both cortical thickness and surface area (Fig. 1b and 1g). As depicted in Fig. 1b, there was a negative correlation between PD-PRS and cortical thickness, implying that the cortex is thinner with higher genetic risk. Conversely, surface area showed a positive correlation with PD-PRS (Fig. 1g). Thus, a dissociation exists between the effects of genetic risk for PD on cortical thickness and surface area consistent with the previously reported low genetic and phenotypic correlation between thickness and surface area ^39^.

**Fig. 1:**
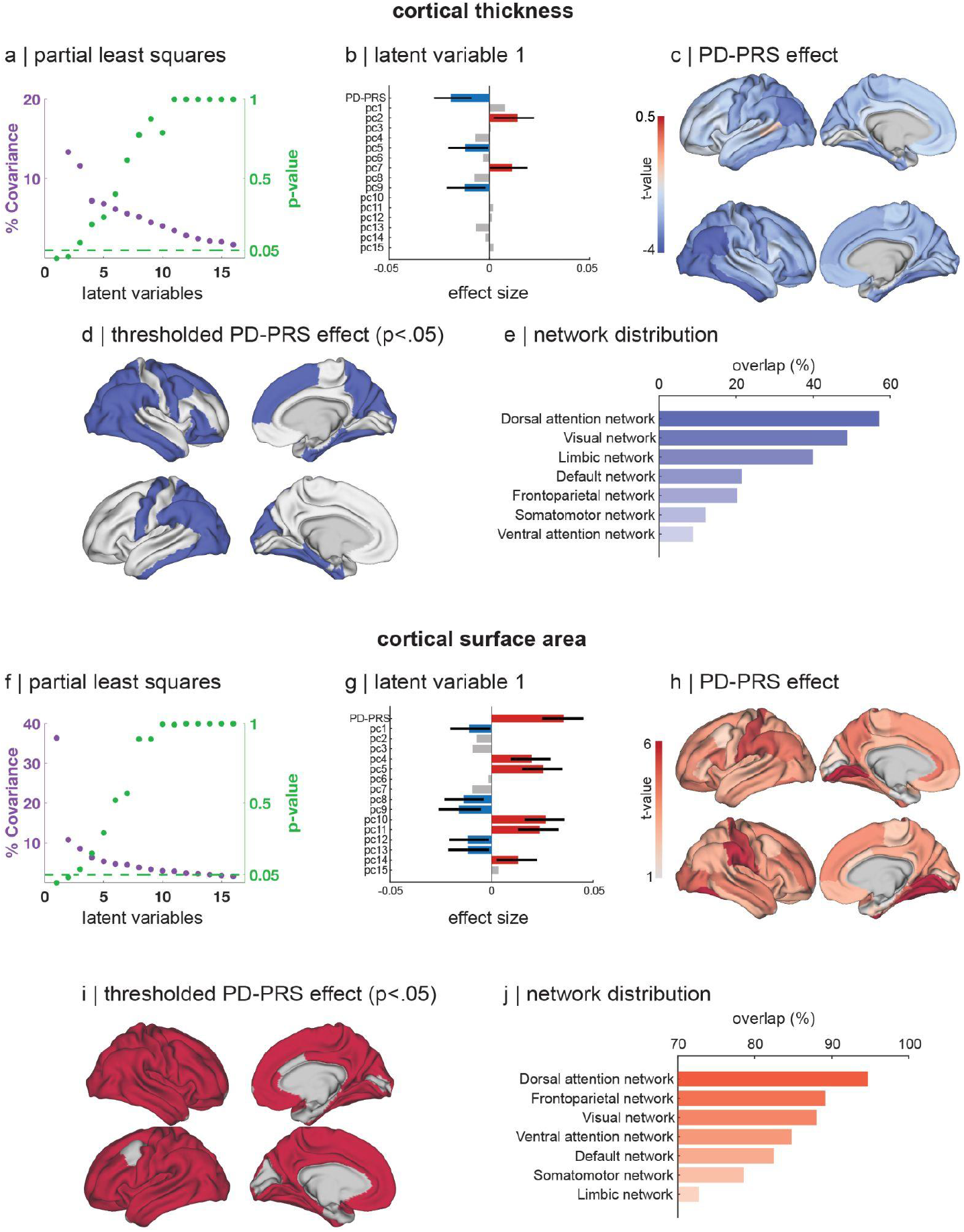
Association between PD-PRS and cortical thickness and surface area. **(a, f)** Covariance explained and permutation p-values for latent variables identified from the partial least squares analysis between genetic components and thickness and surface area, respectively. PC1-PC15 are the first 15 principal components reflecting population stratification (**b, g)** Effect size estimates of the first latent variable from partial least squares analysis. Confidence intervals are calculated by bootstrap testing. (**c, h)** T-statistic maps of the cortical effects of PD-PRS. (**d, i)** Parcels with significant effect of PD-PRS corrected for multiple comparisons. (**e, j)** Distribution of PD-PRS effects in the 7 Yeo intrinsic networks for thickness and surface area, respectively. PD-PRS: Parkinson ‘s Disease Polygenic Risk Score.

Next, to localize the influence of PD-PRS on cortical morphology, we fitted general linear regression models for each cortical region of the DK atlas by considering a regional measurement as the dependent and PD-PRS as the independent variable, while adjusting for the previously mentioned confounds (including age) and correcting for multiple comparisons. We then mapped the t-statistic values of the region-wise analyses for thickness and surface area onto the brain surface for visualization. Consistent with the PLS analyses, PD-PRS showed a negative correlation with cortical thickness (Fig. 1c,d) and a positive correlation with surface area (Fig. 1h,i) in several cortical regions. The patterns of variation across the cortex were similar in the two hemispheres for both measurements. The correlations seemed relatively greater in the more posterior parts of the cortex, particularly for cortical thickness. In order to assess functional correspondence of these patterns, we examined their correlation with seven canonical resting-state functional networks ^40^. DK parcels with statistically significant influence of PD-PRS on cortical thickness were predominantly located in the posterior part of the brain (Fig. 1d) and had the maximum overlap with the dorsal attention resting-state network followed by the visual network, and the least overlap with the ventral attention network (Fig. 1e). On the other hand, the pattern of significant correlation between surface area and PD-PRS was more widespread, covering almost the entire cortical mantle (Fig. 1i) and also having the maximal overlap with the dorsal attention network (Fig. 1j). Effect sizes and statistics for each brain parcel are in the supplementary materials.

### Comparison to Parkinson ‘s disease atrophy distribution

Next, we asked whether the morphometric effects of genetic risk for PD looked like patterns of brain atrophy described in people diagnosed with PD. To answer this, we leveraged findings of two recently published, large-scale investigations of PD-specific cortical alterations ^8,41^. First, we obtained the PD-specific pattern of atrophy by performing deformation-based morphometry (DBM) in a large sample of PD patients in comparison to healthy controls ^42^, as described previously ^41^. DBM is a measure of the change (expansion or atrophy) in the shape of local brain tissue. Prior studies have shown DBM to be a sensitive measure of brain atrophy in PD ^4,43^. Hence, we calculated longitudinal changes via DBM in Parkinson ‘s disease after 2 and 4 years of follow up relative to healthy controls. We then performed correlation tests between PD DBM progression maps and our PD-PRS cortical influence maps for thickness and surface area. We used the “spin-tests” method to generate null models that account for the spatial autocorrelation of cortical brain measurements ^44^. We observed a significant spatial overlap between the PD-PRS influence on cortical thickness and the PD-specific longitudinal pattern of gray-matter atrophy, at 2 years (r = 0.37, p_fdr-spin_ < 0.005) and 4 years (r = 0.37, p_fdr-spin_ < 0.05) (Fig. 2a). In other words, the PD-PRS-related pattern of cortical thickness in healthy participants was found to be spatially correlated with the pattern of cortical thinning in early and more advanced PD patients. We did not observe a correspondence between the PD-PRS effects on cortical surface area and brain atrophy in PD.

**Fig. 2:**
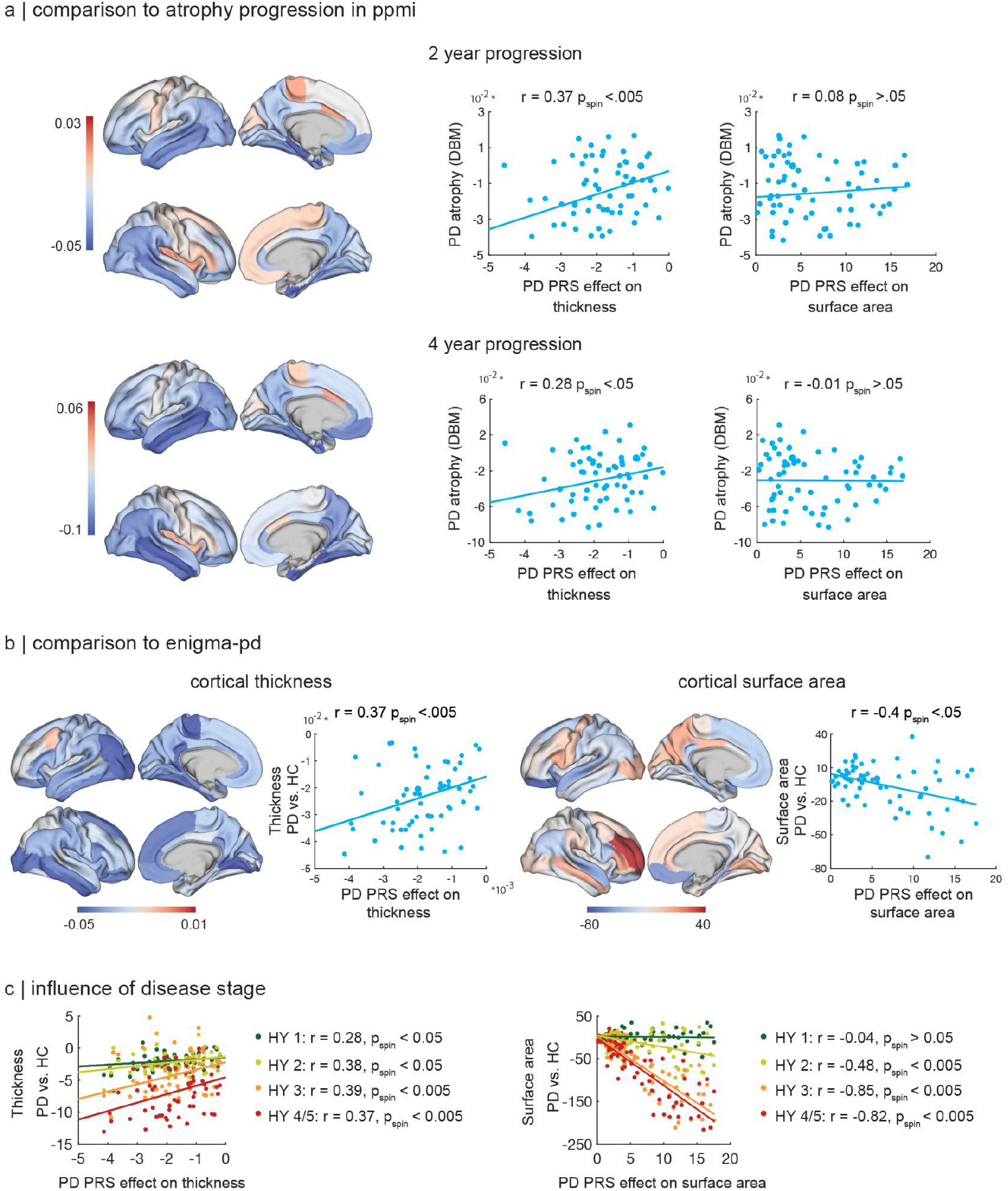
Comparison of the spatial distribution of the PD-PRS effect versus atrophy in Parkinson ‘s disease. Correlation of PD-PRS influence maps for thickness and surface area with: (**a)** PD DBM progression maps after 2 years and 4 years of follow-up from PPMI, and (**b**) cortical thickness and surface area from ENIGMA (PD minus controls). (**c**) Effect of Hoehn and Yahr stage on the correlations in panel b. Brain maps present region-wise correlation coefficient estimates. DBM: deformation-based morphometry; PPMI: Parkinson Progression Markers Initiative; r: correlation coefficient; p_spin_: p-values obtained after multiple comparison correction and spin tests; HY: Hoehn & Yahr stage.

We also compared the PRS effect maps to those of the ENIGMA consortium, who measured cortical thickness and surface area from T1 MRI scans of 2,367 PD patients and 1,183 healthy controls ^8^. We observed a positive correlation between the two cortical thickness maps (r = 0.37, pspin <0.005, Fig. 2b), meaning that the spatial patterns of reduced cortical thickness related to PD-PRS and to PD itself overlapped. We also observed a negative correlation between the two maps of cortical surface area (r = -0.4, pspin <0.05, Fig. 2b). This implies that increased cortical surface area related to the PD-PRS maps onto reduced surface area in PD. Interestingly, for both correlations, the effect size increased with greater disease severity (Fig 2c).

### Role of connectivity

In a subsequent analysis, we tested the propagation model of PD, according to which the neurodegenerative process is mediated by neuronal connectivity. We previously found that the atrophy pattern in PD could be explained by connectivity ^4,24^, meaning that inter-connected regions tend to covary in their degree of volume loss. We now asked whether white-matter connectivity (measured using diffusion MRI) also influences the PD-PRS related pattern of cortical thickness variability. We observed a significant correlation between regional structural connectivity and the influences of PD-PRS on cortical thickness (r = 0.52, p_fdr-spin_ < 0.001) and surface area (r = 0.9, p_fdr-spin_ < 0.001), while taking spatial autocorrelation into account (Fig. 3a). These correlations imply that interconnected areas tend to have similar PD-PRS related influences on their cortical morphometry.

**Fig. 3:**
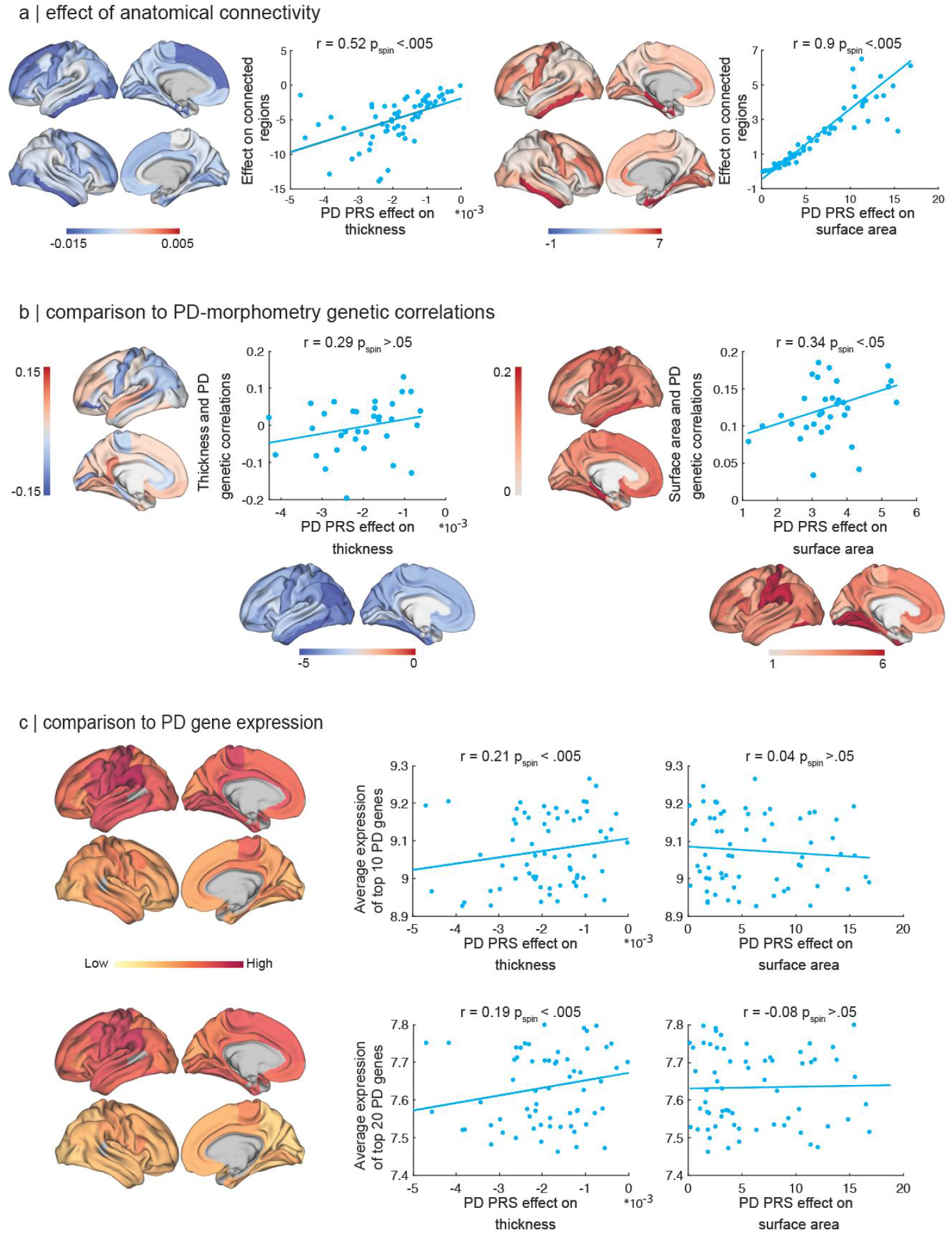
Comparison of PD-PRS effect with other cortical maps. Correlation of PD-PRS influence maps for thickness and surface area with: (**a**) the same measure in physically connected regions (based on diffusion tractography), (**b**) maps of genetic correlations between cortical thickness/area and PD ^45^, and (**c**) cortical expression patterns of the genes near the PD top ten (top row) and top twenty (bottom row) most significant single nucleotide polymorphisms of the GWAS. Gene expression maps derived from the Allen Human Brain Atlas. r = correlation coefficient; p_spin_ = p-values obtained after multiple comparison correction and spin test.

### Comparison to genetic effects on cortical structure

We then sought to compare the cortical patterns found here with recent descriptions of genetic influence on cortical structure. We made use of a recent GWAS of cortical thickness and surface area in 33,992 individuals ^45^. This study calculated genetic correlations between cortical structure and several traits, including PD. We compared our maps of cortical effects of PD-PRS with the maps of genetic correlation between PD risk and cortical thickness and surface area. We averaged cortical values across left and right hemispheres to provide aggregate maps, as was done in the above-mentioned meta-analysis, and then repeated our prior regression models to obtain the PD-PRS influences on these averaged measurements. We next performed correlation analyses between the corresponding t-statistic maps from the meta-analyses and the newly generated maps of PD-PRS influences, while controlling for spatial autocorrelation and multiple comparisons.

There was a significant spatial resemblance between the PD-PRS influence on cortical surface area and its shared genetic effects with PD (r = 0.32, p_fdr-spin_ < 0.05) (Fig. 3a). For cortical thickness there was a marginal but non-significant correspondence between the two maps (r = 0.25, p_fdr-spin_ > 0.05). Note however that only cortical surface area, and not thickness, was found to have a shared genetic effect with PD ^45^.

### Comparison to gene expression maps

We then investigated the cortical expression patterns of the genes near the PD top ten and top twenty most significant single nucleotide polymorphisms (SNP), including *SNCA*, from the PD GWAS study ^11^. For this purpose, we incorporated gene expression data from the Allen Human Brain Atlas ^46^. This approach revealed that PD-PRS influences on cortical thickness, but not surface area, overlap with the cortical expression of PD-specific genes (top ten genes r = 0.21, p_fdr-spin_ < 0.05 - top twenty genes r = 0.19, p_fdr-spin_ < 0.05) (Fig. 3b).

### Virtual histology

Neurodegenerative disorders have been shown to differentially affect certain cell types. Therefore, we investigated if PD-PRS thickness and surface area maps generated here were associated with the prevalence of specific cell types in the cortex by using a virtual histology approach (see details in Methods and Materials). Four cell types showed an association with both PD-PRS related thickness and surface area maps (Fig. 4). Positive correlations were found between the prevalence of excitatory neurons and PRS effects on cortical thickness (r = 0.27, p_fdr_ < 0.05) and surface area (r = 0.26, p_fdr_ < 0.05). Significant negative correlations were also observed between PRS effects and the prevalence of microglia (thickness: r = -0.32, p_fdr_ < 0.05; surface area: r = -0.31, p_fdr_ < 0.05), oligodendrocyte precursors (thickness: r = -0.41, p_fdr_ < 0.05; surface area: r = -0.39, p_fdr_ < 0.05) and astrocytes (thickness: r = -0.43, p_fdr_ < 0.01). No significant correlation was found for the three other cell types (inhibitory neurons, oligodendrocytes, and endothelial cells).

**Fig. 4:**
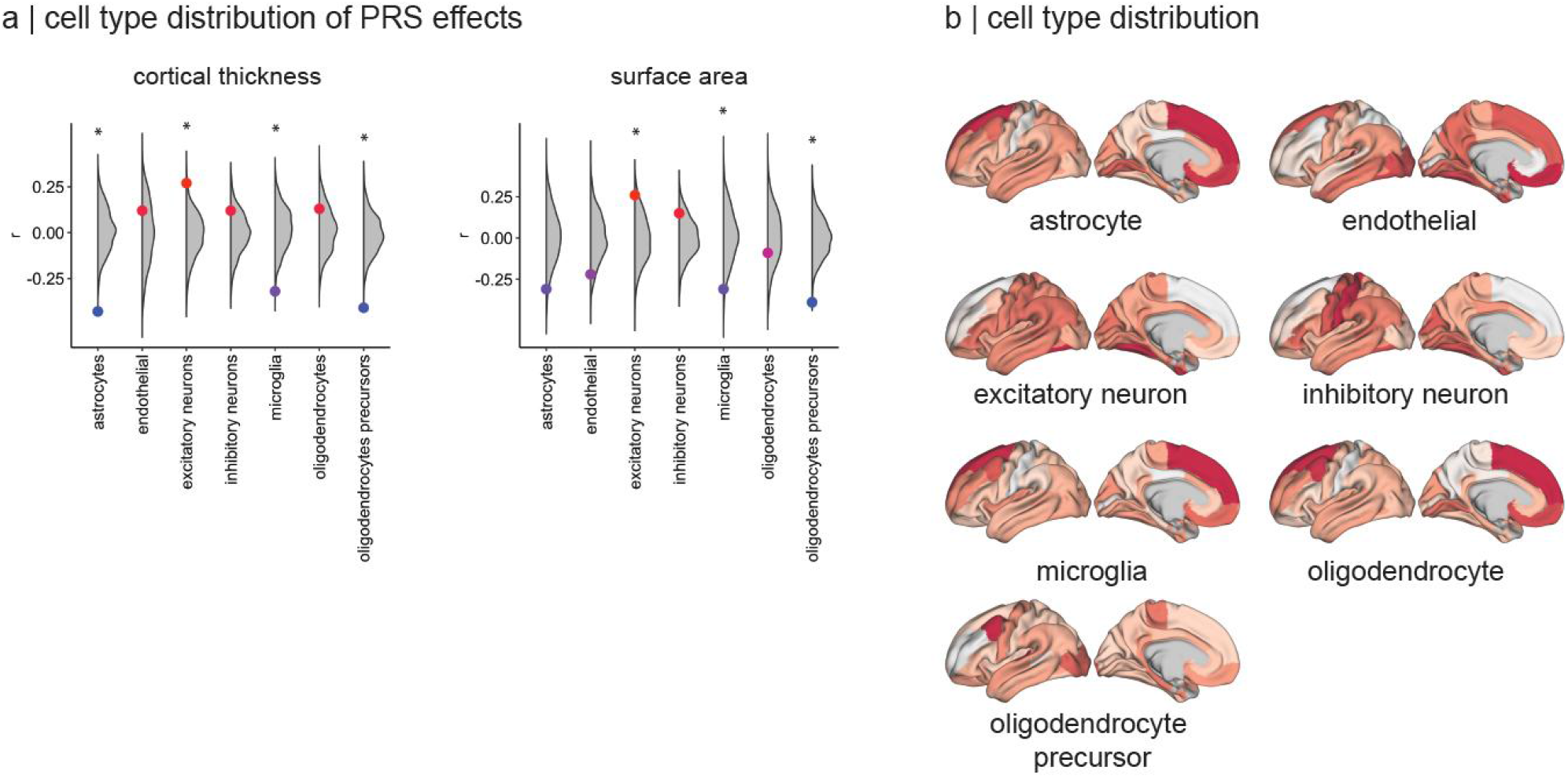
Relation between PD-PRS effect and cortical histological cell type distribution. **(a)** Correlation of PD-PRS influence maps for thickness and surface area with prevalence of specific cell types in the cortex. **(b)** Cell type distribution maps derived from Seidlitz et al. (2020). r = correlation coefficient. * = significant correlation after FDR multiple comparison correction (p < 0.05).

### Gene ontology analysis

Relatedly, to determine the functions of the genes whose expression was spatially associated with PD-PRS related thickness or surface area maps, a gene ontology (GO) enrichment analysis was done. To ensure that the results were not due to the choice of a particular classification system, two platforms, GOrilla and PANTHER, were used (see Methods and Materials). No significant results were associated with surface area. The results associated with cortical thickness from each platform implicated terms related to neuronal signaling. **Table 1** presents the significant GO terms from the genes negatively associated with the PD-PRS thickness map using the GOrilla (n=4) and PANTHER (n=5) platforms, and their fold enrichment. The GO enrichment analysis in both platforms revealed processes related to the cell surface receptor signaling pathways (fold enrichment = 1.40 (GOrilla) and 1.44 (PANTHER)). In addition, processes related to regulation of signaling (fold enrichment = 1.27), signal transduction (fold enrichment = 1.26) and cell communication (fold enrichment = 1.25) from the same set of genes were revealed, using either platform. In other words, cortical regions with more synaptic and signaling activity seem to have greater PD-PRS related cortical thinning. We did not observe a significant GO term for the genes that were positively associated with cortical thickness with either platform. Collectively, these results lend support to the notion that, as with the disease itself, the pattern of PD-PRS related cortical alteration is predominantly associated with synaptic transmission and signaling pathways.

**Table 1.**
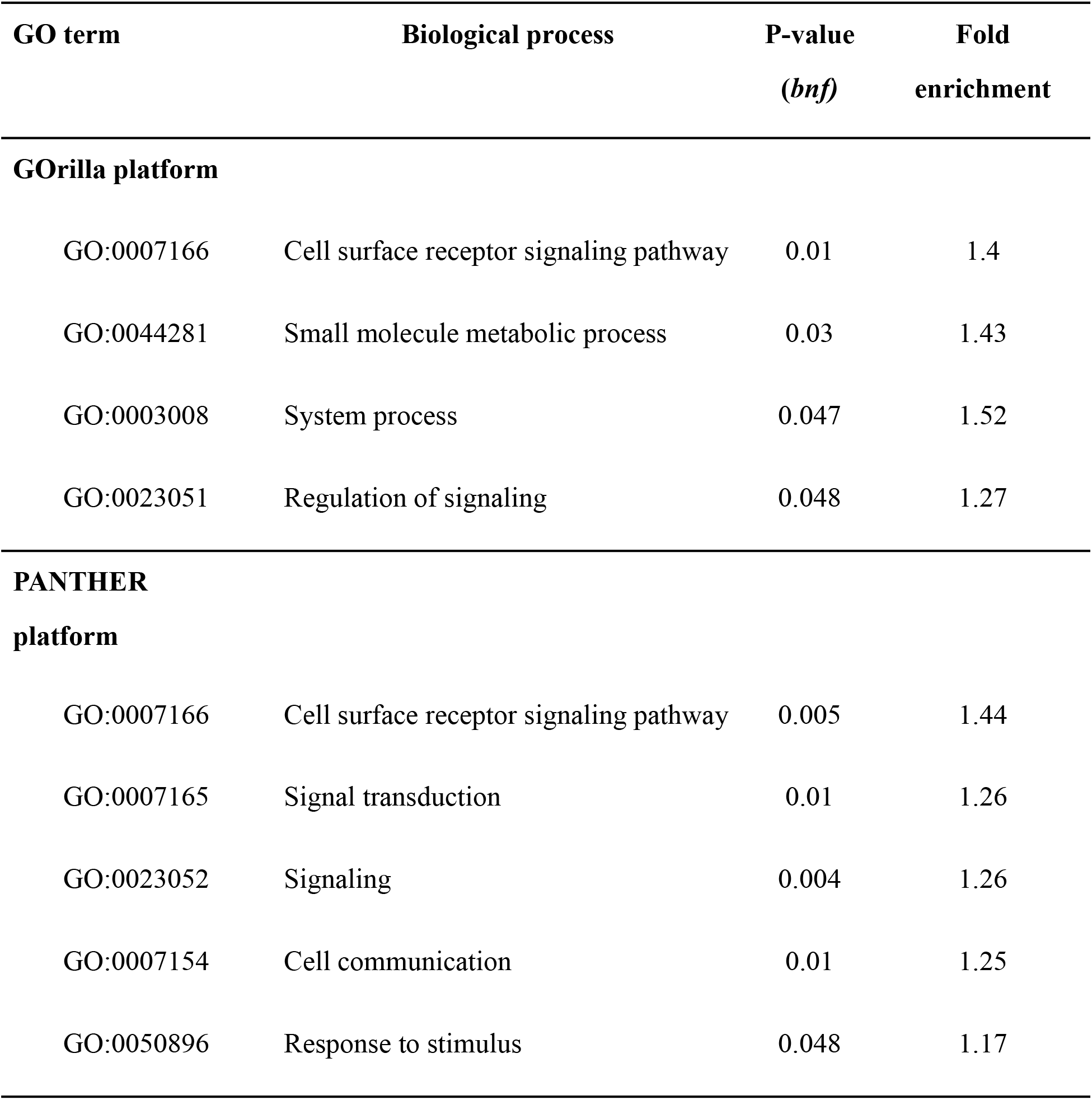
Gene ontology (GO) enrichment analysis of the genes associated with PD-PRS related cortical thinning.

### Behavioral correlates of genetic risk for PD

We exploited the availability of a number of behavioral measures, from a larger sample of UKB to assess their relationship with the PD-PRS (Table 2, Fig. 5a). We found significant negative associations between PD-PRS and smoking (packs/year, n = 134,861, *t* = -3.48, p_fdr_ < 0.005), frequency of alcohol consumption (n = 142,516, *t* = -4.08, p_fdr_ < 0.005), body mass index (n = 448,461, *t* = -4.01, p_fdr_ < 0.005), and sleep duration (n = 446,714, *t* = 2.77), supporting the notion that behavioral alterations seen in PD may also develop in healthy subjects with genetic susceptibility to PD prior to or in the absence of overt parkinsonism. There was a positive association between PD-PRS and sleep duration (n = 446,714, *t* = 2.77, p_fdr_ < 0.01). Coffee consumption (n = 448,360, *t* = 1.04), educational attainment (n = 449,137, *t* = 1.53) and fluid intelligence (n = 147,365, *t* = 1.01) did not show a statistically significant association.

**Table 2.** Behavioral correlates of genetic risk for PD. N; sample size for the analysis. fdr: false discovery rate corrected p-value.

| **Behavior/Habit** | **N** | **T-value** | **P-value**<br>**(*fdr*)** |
| --- | --- | --- | --- |
| **Smoking cigarette**<br>Pack/year | 134,861 | -3.48 | **0.001** |
| **Alcohol intake frequency**<br>Daily or almost daily<br>Three or four times a week<br>Once or twice a week<br>Three times a month<br>Social occasions only<br>Never | 142,516 | -4.08 | **2.07e-04** |
| **Coffee intake**<br>Cups/day | 448,360 | 1.04 | 0.31 |
| **Sleep duration**<br>Hours/day | 446,714 | 2.77 | **0.01** |
| **Body mass index**<br>Kg/m<sup>2</sup> | 448,461 | -4.01 | **2.07e-04** |
| **Educational attainment**<br>College or university degree<br>A levels/AS levels or equivalent<br>O levels/GCSEs or equivalent<br>CSEs or equivalent<br>NVQ or HND or HNC or equivalent<br>Other professional qualifications e.g., nursing,<br>teaching<br>None of the above | 449,137 | 1.53 | 0.17 |
| **Fluid intelligence**<br>Number of correct answers given to the fluid<br>intelligence questions | 147,365 | 1.01 | 0.31 |

**Fig. 5:**
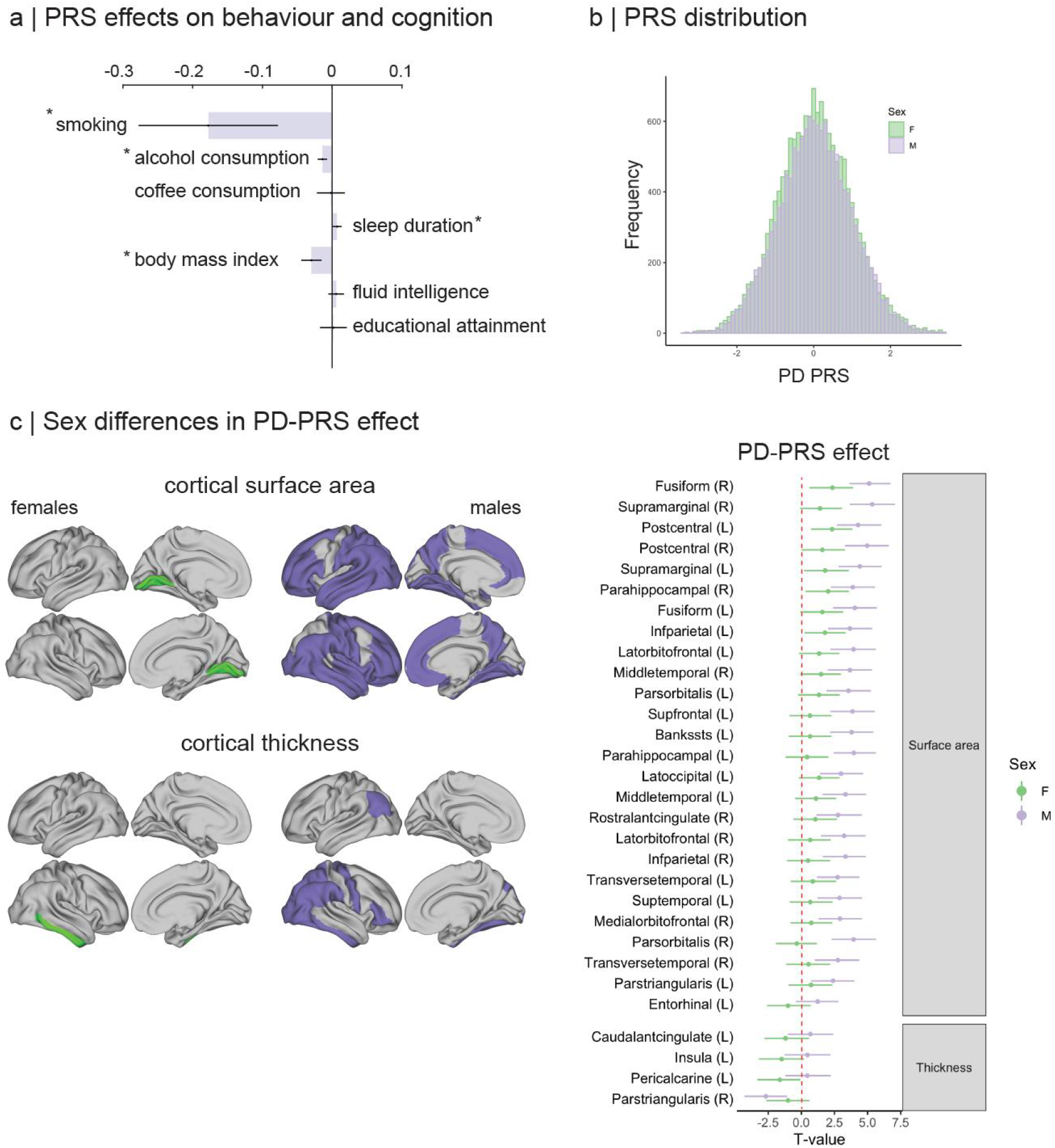
Phenotypic and sex effects. **(a)** Correlation between PD-PRS and characteristic PD phenotypes in healthy participants of the UK Biobank (see Table 2). * = significant correlation after multiple comparison correction (i.e., corrected p-value < 0.05). **(b)** Distribution of the PD-PRS in males (n=13,976) and females (n=15,125) in this study. (**c**) Thresholded maps (p<.05 corrected for multiple comparisons) and regional T-values with 95% confidence intervals of the significant correlations between PD-PRS and cortical measures of surface area and thickness for female and male participants.

### Sex differences

PD affects males more than females ^31^ and males have a more severe course and show greater patterns of cortical thinning ^47^ We sought to understand whether this was related to differences in genetic risk. First, we found that the distribution of polygenic risk was identical for males and females (Fig. 5b). However, the cortical thinning and expanded surface area associated with PD-PRS were essentially only observed in males (Fig. 5c). There were no significant differences for cortical thickness; however, for surface area, most parcels showed a statistically significant difference, with a greater effect size in males. Indeed, the 95% confidence intervals show that the widespread increase in cortical surface area associated with PD-PRS was only present in males, except for one region also showing the effect in females.

## Discussion

We present an investigation of the neural correlates of genetic risk for PD using neuroimaging and genotyping data from 29,101 healthy older individuals. Higher genetic risk was associated with lower cortical thickness in mostly posterior areas but greater cortical surface area globally. A similar dichotomy has also been reported in PD (i.e. increased area, reduced thickness), albeit in a more spatially restricted pattern and in studies with relatively small sample sizes ^48,49^. Measures of cortical surface area and thickness from T1-weighted MRI are independent: while both are heritable, they demonstrate little genetic overlap and are thought to result from different neuro-developmental processes ^39,45,50^. This has been linked to the radial-unit hypothesis ^51^, wherein neural progenitor differentiation in early embryogenesis is reflected in the number of neocortical columns and hence surface area, while events later in development influence the number of neurons and synapses per column and are reflected in cortical thickness ^52^. In later life, age or disease-related neurodegeneration result in reductions in cortical thickness ^53^. A GWAS study of cortical morphometry confirmed the ontogenetic dichotomy between thickness and surface area ^45^: both anatomical features were related to genetic regulatory sites, but surface area was associated with elements active in the mid-fetal period of development while thickness was mostly linked to regulatory activity in adulthood. This suggests that the cortical thickness and surface area associations found here may represent different manifestations of genetic risk for PD.

The regions showing PD-PRS-related cortical thinning were mostly in occipital and parietal lobes, as well as medial and orbital prefrontal areas (Fig. 1). This pattern overlapped significantly with the cortical thinning distribution seen in PD in two large cohorts: ENIGMA ^8^ and PPMI ^41,42^. The associations between PD-PRS and PD cortical thinning were stronger with advanced disease stage (from the ENIGMA study) and matched progressive cortical atrophy patterns after 4 years (in the PPMI dataset). Thus, the genetically-determined cortical thinning pattern corresponds spatially to the progressive neural tissue loss seen in PD: areas showing lower cortical thickness in people with higher genetic risk are the ones that tend to atrophy faster in PD.

The PD-PRS-associated cortical thinning pattern also overlapped with the cortical expression of the most significant associated genes from the GWAS ^11^. Overlap between patterns of cortical thickness and expression of genes that influence cortical thickness has also been described for normal cortical development ^51^ and several neurodevelopmental disorders ^54^. Twelve of the top 20 genes from the PD GWAS are associated with the autophagy-lysosomal pathway implicated in protein homeostasis and alpha-synuclein accumulation (Table 3) ^13,14^. Their greater expression in areas affected by the PD-PRS could represent a neurodevelopmental influence, but it could also represent accelerated age-related synaptic loss in higher-risk individuals. We note however, that the function of many genes contributing to the PD GWAS remains unknown.

**Table 3.** Top genes from the PD GWAS, taken from Table S2 of Nalls et al. (2019). Rows in red indicate genes whose function is related to the autophagy-lysosomal pathway. Duplicate genes in light gray. SNP: single nucleotide polymorphism. QTL: quantitative trait locus.

| SNP | Nearest Gene | QTL Nominated Gene | P, all studies |
| --- | --- | --- | --- |
| rs356182 | SNCA | SNCA | 3.89E-154 |
| rs356203 | SNCA |  | 5.16E-149 |
| rs34637584 | LRRK2 | LRRK2 | 3.61E-148 |
| rs35749011 | KRTCAP2 |  | 1.72E-70 |
| rs34311866 | TMEM175 | TMEM175 | 9.98E-70 |
| rs62053943 | CRHR1 | WNT3 | 3.58E-68 |
| rs199453 | NSF |  | 8.62E-67 |
| rs356228 | SNCA |  | 9.53E-49 |
| rs7225002 | KANSL1 |  | 1.84E-40 |
| rs1474055 | STK39 | STK39 | 2.54E-39 |
| rs7221167 | MAPT-AS1 |  | 1.36E-37 |
| rs10847864 | HIP1R | HIP1R | 1.47E-37 |
| rs5019538 | SNCA | SNCA | 1.13E-36 |
| rs10513789 | MCCC1 | MCCC1 | 1.22E-34 |
| rs823118 | NUCKS1 | NUCKS1 | 1.11E-29 |
| rs76904798 | LRRK2 | LRRK2 | 1.52E-28 |
| rs4698412 | BST1 | BST1 | 2.06E-28 |
| rs117896735 | INPP5F | SEC23IP | 2.36E-28 |
| rs112485576 | HLA-DRB5 | HLA-DRB5 | 6.96E-28 |
| rs17686238 | MAP3K14 |  | 8.25E-27 |
| rs199351 | GPNMB | GPNMB | 5.25E-26 |
| rs1867598 | ELOVL7 | ELOVL7 | 2.52E-23 |
| rs12456492 | RIT2 |  | 3.80E-23 |
| rs76763715 | GBAP1 | GBAP1 | 1.59E-22 |

Age-related cortical thinning is thought to be due to loss of neuropil and synaptic density rather than neuronal loss ^55,56^. Virtual histology approaches based on MRI support the theory that cortical thinning with aging or neurodegeneration reflects loss of dendritic arbors and synapses ^41,57,58^. Similarly, postmortem studies show that PD appears to be associated with synaptic loss but normal neuronal numbers in the cortex ^59,60^. Our current results also implicate synaptic loss, as the PD-PRS related pattern of cortical thinning overlapped spatially with the expression of genes involved in synaptic and signaling activities.

Thus, an intriguing possibility is that lower cortical thickness associated with genetic risk for PD in older adults shown here may represent synaptic loss similar to that seen in PD, but insufficient to cause neurological symptoms. Indeed, autosomal-lysosomal pathway dysfunction leads to imapired protein homeostasis and alpha-synuclein accumulation at the synapse ^61^. It is possible that high genetic risk for PD derives in part from an enhanced susceptibility to lysosomal dysfunction, toxic protein accumulation, and synaptic damage in older age, and that these phenomena occur in people who do not have overt PD.

In PD, the spatial pattern of cortical atrophy follows anatomical connectivity ^4,23^, which has been hypothesized to reflect neuronal propagation of alpha-synuclein misfolding ^21,24^. Therefore, if the cortical thickness pattern associated with PD-PRS reflects reduced synaptic density due to similar mechanisms, it may be expected to also follow brain connectivity. Indeed, this is what we found: the PD-PRS effect on cortical thickness in any region was proportional to the summed effect in its connected neighbors.

It should be noted that there is no evidence that healthy adults with higher genetic risk for PD have a subclinical form of the disease or harbor any PD-like pathology. Nonetheless, it is interesting to note that a population postmortem study in elderly individuals not diagnosed with PD identified Lewy pathology in a distribution that resembled PD in 41% of individual brains ^62^.

The finding of diffusely increased cortical surface area with higher PD-PRS is in contrast to the reduced cortical thickness. As mentioned earlier, a GWAS study of cortical morphometry in 33,992 individuals supported the theory of divergent genetic contributions to cortical surface area and thickness ^45^. These authors also found a positive genetic correlation between total cortical surface area and three phenotypes: PD, educational attainment, and cognitive ability. This may be consistent with studies that have identified intelligence and educational attainment as risk factors for PD ^27,63^, although in our sample there was no effect of PD-PRS on intelligence or educational attainment. It is not clear why the PD-PRS is associated with increased cortical surface area, but the finding raises the possibility that some of the genetic risk for PD derives from effects on neural progenitors and a consequent increase in cortical surface area. Thus, the brain-based genetic vulnerability to PD might be present from birth. Why a brain with greater cortical surface area, and hence neuronal columns, is also vulnerable to PD pathology remains unknown.

We also sought to determine whether people with high PD-PRS displayed neurobehavioral phenotypes seen in PD. We found that, in the larger UKB sample (Ns=140,000 to 400,000), PD-PRS is associated with a reduced tendency to smoke or drink alcohol, lower BMI, and greater sleep duration. These associations are also consistently described in PD ^11,26,29,64^. While some research has proposed a protective effect of cigarette smoking on the brain, the association found here is more compatible with the opposite causality - namely that genetic predisposition to PD is associated with a reduced tendency for behaviors that depend on reinforcement, namely cigarette and alcohol use and over-eating. There was also a positive association between PD-PRS and questionnaire-derived sleep duration, which has also been described in PD ^64^. Intelligence and educational attainment did not show a statistically significant association with genetic risk.

Finally, we looked for sex differences in the manifestations of genetic risk. The higher incidence and severity of PD in men has been variously attributed to sex differences in expression of disease-related genes ^65^ and protective effects of estrogen on dopamine neuronal death and on alpha-synuclein accumulation ^66,67^. This could account for the fact that PD-PRS related cortical thinning was seen in men only, however the male-female difference in cortical thickness was not statistically significant. On the other hand, the increased cortical surface area was only seen in men and the difference was statistically significant. If the genetic effects on cortical surface are indeed active during fetal development ^45,51^, this would suggest that sex differences in PD are partly attributable to genetically determined lifelong differences in brain morphometry. However, little is known about sex differences in genetic effects on cortical development.

There are some limitations to this study. The PD-PRS only accounts for 16-36% of the heritability of PD ^11^, meaning that genetic effects not studied here may also contribute to vulnerability to PD in different ways. Also, the findings are correlative and do not prove a causal relationship between either of the morphometric brain patterns uncovered here and the development of PD. Finally, future studies should investigate MRI measures of basal ganglia and white matter integrity, which are also available in UKB. Nonetheless, our results provide evidence that the genetic risk for PD manifests in the brain of healthy individuals, and that it resembles morphometric changes seen in PD itself. We hope that these findings may be of use in understanding and modeling prodromal PD, and eventually developing neuroprotective interventions.

## Materials and Methods

### Data resource

UK Biobank (UKB) is a large prospective study, covering half a million participants aged between 40-69 at the baseline assessment period (2006-2010). The cohort is a shared resource to promote research into etiology and mechanisms of a wide range of health-related conditions ^25^. UKB encompasses a comprehensive set of information on lifestyle, environment, medical history, physical measures and biological samples. Our study involved data from a subset of 42,488 participants (final sample 29,101 after exclusions as described below) with brain-imaging measures of cortical morphology including thickness and surface area ^32,35^. This study was conducted under UKB approvals for application #35605 (PI Dagher). Participants provided written, informed consent (http://biobank.ctsu.ox.ac.uk/crystal/field.cgi?id=200). Exclusion criteria for the current analysis were a history of bipolar or any neurological disorder including a comprehensive set of degenerative, vascular, traumatic, infectious brain pathologies; first degree family history of Parkinson ‘s disease; body mass index (BMI) > 35; relation to another participant closer than cousin; genetic and self-reported sex mismatch; and non-European ancestry. The latter criterion is necessary at the present time to ensure valid genetic analyses, as the GWAS studies we used were performed in European ancestry individuals. After all exclusion criteria, the final sample consisted of 29,101 individuals (13,976 males, 15,125 females). This study was approved by the McGill University Health Centre Research Ethics Board. Open-access data from other sources derive from studies that were approved by the relevant local ethics boards.

### Data analysis software

The software packages used for analysis of data in this study include FreeSurfer (http://surfer.nmr.mgh.harvard.edu/), FSL (https://fsl.fmrib.ox.ac.uk/fsl/fslwiki/) and PLINK ^68^, available at http://pngu.mgh.harvard.edu/purcell/plink/. The analyzed data were imported into MATLAB (The Mathworks, Inc.) and Python (https://www.python.org/) for further computations. Gene expression maps were generated using abagen (https://github.com/rmarkello/abagen).

### Brain imaging and preprocessing procedures

The structural magnetic resonance imaging data were acquired as high-resolution T1-weighted images using a 3D MPRAGE sequence at 1-mm isotropic resolution at three imaging sites in the United Kingdom with identical scanners and acquisition protocols. Data were submitted to automated preprocessing and quality control pipelines ^35^. For the current analysis, we used cortical thickness and surface area values generated with FreeSurfer and parcellation of the surface using the DK atlas, available in Freesurfer. Data from 42,488 participants of the UKB data release in early 2021 were downloaded. We included data from 29,101 individuals, after applying the exclusion criteria mentioned earlier.

### PD-PRS calculation

We consider the 487,410 samples included in the 2019 release of UKB. Of the samples, 31,386 are included among the 42,488 samples in UKB for which brain imaging data are available, after the genotyping quality control procedures for sample removal ^69^. As noted above, we excluded subjects with non-European ancestry based on self-reported ancestry and genetic principal component thresholds. In addition, we excluded individuals based on relatedness, closer than cousins, creating a maximally unrelated study sample. Subjects whose self-reported sex information did not match the genotyping were also excluded. The sample size after these exclusions was 31,386. We then excluded first-degree relatives of people with PD leaving a final sample of 29,101. The PD-PRS was calculated using the effect size of 1805 SNPs from the latest PD GWAS summary statistics ^11^ using PRSice-2 ^33^ without pruning or thresholding. These two steps were omitted because the 1805 SNPs were tested by Nalls et al. in a discovery cohort and replicated. Note also that the sample for this GWAS also included 18,618 first-degree relatives of people with Parkinson ‘s Disease in the UKB, as proxy cases, and that these individuals are excluded from the present analysis.

### Confounds

The following comprehensive set of imaging and genetic confounds was used in the analyses: age, age squared, sex, head motion during functional MRI, scan date, site and its interactions with the other confounds ^36^, the top 15 population genetic principal components (explaining most of the data variance) supplied by UKB, and genotyping batch.

### Partial least squares analysis

Partial least squares (PLS) regression is a multivariate method used for finding the relations between two sets of variables ^38,70^. The analysis tries to find linear combinations of the input features that maximally covary with each other. Here, the two variable sets were cortical thickness and surface area, on the one hand, and genetic components including the PD-PRS and 15 top genetic ancestry components, on the other. This analysis was performed to determine whether a combination of genetic factors, and in particular, the PD-PRS, can explain any degree of variation in the brain cortical measurements. For this purpose, we first regressed out imaging-related confounds (described above) from the brain MRI measurements. Singular value decomposition was then applied to the correlation matrix between the brain and genetic data. We used permutation tests with 500 repetitions to determine the significance of the covariance explained for each latent variable. We then used the bootstrapping method (i.e., random resampling with replacement, *n* = 500 times) to calculate the confidence interval for individual coefficients for each variable loaded in a given latent variable ^38^.

### Linear regression models

Our second analysis involved performing several regression models for each cortical measurement separately, in order to localise the spatial relationship between PD-PRS and cortical surface area and thickness. We again included all the confounders mentioned earlier as covariates in our regression models. P-values were then corrected for multiple comparisons using the false discovery rate (FDR) approach over the number of brain measures. We took p=0.05 (corrected) as the significance threshold.

### Correspondences between two cortical maps

To identify correspondences between the topographies of any two cortical maps, we performed correlation tests. As most standard methods for statistical inference do not account for spatial properties of the underlying brain maps, we used a spherical projection null model, or spin-test, that permutes cortical regions and generates null distributions while preserving spatial autocorrelation ^44^. This model overcomes data loss caused by rotation of the medial wall (containing no data) into the cortical surface by assigning the nearest data to the missing parcels. The statistical significance of each test was assessed and reported against the null distributions from 1,000 repetitions of the spin test (i.e., p_spin_).

### Connectivity Analysis

To test whether cortical measures in any area were influenced by the same measure in connected neighbors we used structural connectivity data from diffusion MRI in an independent sample of 70 healthy participants ^71^, as described previously ^41,72^. A deterministic connectivity matrix was built from the normalized number of streamlines between each region pair divided by the average length of the streamlines and the surface area of the two regions. Correlations were computed between the cortical thickness or surface area in each region and its collective neighborhood thickness or surface area, defined as the mean value in all connected regions divided by the number of connected regions. Statistical significance was tested against a null model preserving spatial autocorrelation, as described in the previous paragraph.

### Cell type analysis: virtual histology

We investigated if spatial patterns of PD-PRS-related cortical thickness and surface area effects were associated with the relative distribution of specific cell types in the cortex, notably astrocytes, endothelial cells, microglia, excitatory and inhibitory neurons, oligodendrocytes and oligodendrocyte precursors ^73^. Each cell type was associated with its corresponding gene list, as derived by Seidlitz et al. ^54^ from post-mortem single-cell RNA sequencing studies of human cortical samples. The spatial patterns of expression of the genes on each list were then computed using post-mortem gene-expression data from the Allen Human Brain Atlas ^46^ with the *abagen* toolbox (https://github.com/rmarkello/abagen) ^74,75^. Pearson ‘s correlations were calculated between the thickness or surface area measurement of each region of PD-PRS-related cortical maps and the region ‘s average gene expression of each cell class. All the correlations were corrected for multiple comparisons and were also tested against the nulls obtained from the spin test (p_fdr-spin_) (Vázquez-Rodríguez et al., 2019). The protocol used here is detailed in Hansen et al. ^76^.

### Gene expression and gene ontology enrichment analysis

We selected the top ten and top twenty most influential genes in the meta-analysis of PD GWAS ^11^. The selected genes were those in closest proximity to single nucleotide polymorphisms with the highest contribution in the GWAS, determined on the basis of p-value. We then generated cortical expression maps for these genes by using the gene expression data from the Allen Human Brain Atlas ^46^ and the *abagen* toolbox ^75^. The Allen atlas consists of microarray gene expression measurements from 6 donor brains sampled at 500 sites per hemisphere. We compared these spatial gene expression patterns to the PD-PRS related patterns while controlling for spatial autocorrelation ^44^.

Separately, a gene ontology (GO) enrichment analysis was performed to explore the biological processes related to gene expression from the spatial patterns of PD-PRS influence on cortical thickness and surface area. We extracted the average gene expression value for all genes available in the Allen Human Brain Atlas genetics dataset for each of the cortical regions of the DK atlas using the *abagen* toolbox. We only retained the genes whose expression significantly correlated with the pattern of PD-PRS related cortical measures after the FDR and spatial auto-correlation corrections. These yielded lists of genes whose expression pattern was positively or negatively correlated with thickness (positive correlation n = 1,065 genes, negative correlation n = 1,194 genes) and surface area (positive correlation n = 34 genes, negative correlation n = 75 genes) PD-PRS-related maps. We next investigated if the proportion of the GO terms for these genes significantly differed from the proportion of GO terms found for all genes from the dataset. Two gene ontology platforms were used to obtain GO terms, the Gene Ontology enRIchment anaLysis and visuaLizAtion tool (Gorilla) ^77^ and the PANTHER Classification System ^78^. Of the 15,633 genes available in the Allen Human Brain Atlas genetics dataset, 13,992 and 14,657 genes were associated with a GO term in the GOrilla (GO Process) and PANTHER (GO biological process) platforms, respectively. Supported gene IDs are available from the Gorilla (http://cbl-gorilla.cs.technion.ac.il) and PANTHER (www.pantherdb.org) websites. For both platforms, a statistical over-representation analysis was conducted with Bonferroni correction to control for multiple comparisons. Whereas a hypergeometric model was implemented in GOrilla, the Fisher ‘s Exact test was used in PANTHER.

### PD-PRS and PD behavioral characteristics

We asked whether PD-PRS relates to a number of previously identified characteristic phenotypes observed in PD. For this, we included any neurologically healthy participant from UKB with data for the following traits: pack/year of cigarette smoking (if ever done), alcohol intake frequency (daily or almost daily, three or four times a week, once or twice a week, three times a month, social occasions only, or never), coffee intake (number of cups per day), sleep duration (hours of sleeping including daily naps per day), educational attainment (defined as: college or university degree, A levels/AS levels or equivalent, O levels/GCSEs or equivalent, CSEs or equivalent, NVQ or HND or HNC or equivalent, other professional qualifications, e.g., nursing, teaching, or none), measured body mass index (BMI; kg/m^2^) and measured fluid intelligence (the capacity to solve problems that require logic and reasoning ability, independent of acquired knowledge). Subjects with a positive history of neurological or psychiatric disorders (as listed earlier in our exclusion criteria) and/or family history of first-degree PD relatives were excluded from this analysis. We calculated PD-PRS for each participant and then ran linear regression analysis between each behavioral feature and the PD-PRS, while controlling for the effect of age, sex, and the first 15 genetic ancestry principal components. FDR correction was further applied on the p-values to correct for multiple comparisons. Sample sizes for this analysis varied between 130,000 and 450,000 and are listed for each measure in the results section.

### Sex differences

We first compared the distribution of PD-PRS in males (n=13,976) and females (n=15,125) and confirmed that it was identical (Fig. 5b). We then computed the relationship between PD-PRS and cortical thickness and surface area for each group, exactly as described above. To compare the t-stats of the PD-PRS effect on cortical maps between males and females, 95% confidence intervals and significance level were derived from bootstrapping (n = 1000 times; with replacement - in order to create a null distribution) of the male and female samples separately and re-running the linear regression iteratively for each brain parcel.

## Data Availability

All data produced in the present study are available upon reasonable request to the authors

https://www.ukbiobank.ac.uk/

https://www.ppmi-info.org/

https://alleninstitute.org/

## Data Availability

The brain maps of cortical thickness and surface area correlation with PD-PRS will be made available upon request.

## Acknowledgements

This work was funded by grants from the Canadian Institutes of Health Research, the Michael J Fox Foundation for Parkinson ‘s Research, the Alzheimer ‘s Association, the Weston Brain Institute, and the Healthy Brains for Healthy Lives (HBHL) initiative of McGill University. NA received a scholarship from the Montreal Neurological Institute.

We thank Ysbrand van der Werf and Max Laansma for sharing the Enigma maps and for comments on the manuscript.

## References

1. Berg, D. et al. Prodromal Parkinson disease subtypes - key to understanding heterogeneity. Nat. Rev. Neurol. 17, 349–361 (2021).

2. Braak, H. et al. Staging of brain pathology related to sporadic Parkinson ‘s disease. Neurobiol. Aging 24, 197–211 (2003).

3. Rahayel, S. et al. A Prodromal Brain-Clinical Pattern of Cognition in Synucleinopathies. Ann. Neurol. 89, 341–357 (2021).

4. Zeighami, Y. et al. Network structure of brain atrophy in de novo Parkinson ‘s disease. eLife vol. 4 Preprint at https://doi.org/10.7554/elife.08440 (2015).

5. Burré, J., Sharma, M. & Südhof, T. C. Cell Biology and Pathophysiology of α-Synuclein. Cold Spring Harb. Perspect. Med. 8, (2018).

6. Brundin, P. & Melki, R. Prying into the Prion Hypothesis for Parkinson ‘s Disease. J. Neurosci. 37, 9808–9818 (2017).

7. Schulz-Schaeffer, W. J. The synaptic pathology of alpha-synuclein aggregation in dementia with Lewy bodies, Parkinson ‘s disease and Parkinson ‘s disease dementia. Acta Neuropathol. 120, 131–143 (2010).

8. Laansma, M. A. et al. International multicenter analysis of brain structure across clinical stages of Parkinson ‘s disease. Mov. Disord. (2021) doi:10.1002/mds.28706.

9. Gcwensa, N. Z., Russell, D. L., Cowell, R. M. & Volpicelli-Daley, L. A. Molecular Mechanisms Underlying Synaptic and Axon Degeneration in Parkinson ‘s Disease. Front. Cell. Neurosci. 15, 626128 (2021).

10. Bellucci, A. et al. Review: Parkinson ‘s disease: from synaptic loss to connectome dysfunction. Neuropathol. Appl. Neurobiol. 42, 77–94 (2016).

11. Nalls, M. A. et al. Identification of novel risk loci, causal insights, and heritable risk for Parkinson ‘s disease: a meta-analysis of genome-wide association studies. Lancet Neurol. 18, 1091–1102 (2019).

12. Gan-Or, Z., Dion, P. A. & Rouleau, G. A. Genetic perspective on the role of the autophagy-lysosome pathway in Parkinson disease. Autophagy 11, 1443–1457 (2015).

13. Senkevich, K. & Gan-Or, Z. Autophagy lysosomal pathway dysfunction in Parkinson ‘s disease; evidence from human genetics. Parkinsonism Relat. Disord. 73, 60–71 (2020).

14. Hou, X., Watzlawik, J. O., Fiesel, F. C. & Springer, W. Autophagy in Parkinson ‘s Disease. J. Mol. Biol. 432, 2651–2672 (2020).

15. Lin, X. et al. Leucine-rich repeat kinase 2 regulates the progression of neuropathology induced by Parkinson ‘s-disease-related mutant alpha-synuclein. Neuron 64, 807–827 (2009).

16. Gan-Or, Z., Liong, C. & Alcalay, R. N. GBA-Associated Parkinson ‘s Disease and Other Synucleinopathies. Curr. Neurol. Neurosci. Rep. 18, 44 (2018).

17. Konno, T., Ross, O. A., Puschmann, A., Dickson, D. W. & Wszolek, Z. K. Autosomal dominant Parkinson ‘s disease caused by SNCA duplications. Parkinsonism Relat. Disord. 22 Suppl 1, S1–6 (2016).

18. Escotts-Price, V. et al. Polygenic risk of Parkinson disease is correlated with disease age at onset. Ann.Neurol. 77, 582–591 (2015).

19. Paul, K. C., Schulz, J., Bronstein, J. M., Lill, C. M. & Ritz, B. R. Association of Polygenic Risk Score With Cognitive Decline and Motor Progression in Parkinson Disease. JAMA Neurol. 75, 360–366 (2018).

20. Henderson, M. X. et al. Spread of α-synuclein pathology through the brain connectome is modulated by selective vulnerability and predicted by network analysis. Nat. Neurosci. 22, 1248–1257 (2019).

21. Luk, K. C. et al. Pathological α-synuclein transmission initiates Parkinson-like neurodegeneration in nontransgenic mice. Science 338, 949–953 (2012).

22. Rahayel, S. et al. Differentially targeted seeding reveals unique pathological alpha-synuclein propagation patterns. Brain (2021) doi:10.1093/brain/awab440.

23. Yau, Y. et al. Network connectivity determines cortical thinning in early Parkinson ‘s disease progression. Nat. Commun. 9, 12 (2018).

24. Zheng, Y.-Q. et al. Local vulnerability and global connectivity jointly shape neurodegenerative disease propagation. PLoS Biol. 17, e3000495 (2019).

25. Sudlow, C. et al. UK biobank: an open access resource for identifying the causes of a wide range of complex diseases of middle and old age. PLoS Med. 12, e1001779 (2015).

26. Dagher, A. & Robbins, T. W. Personality, addiction, dopamine: insights from Parkinson ‘s disease. Neuron 61, 502–510 (2009).

27. Fardell, C., Torén, K., Schiöler, L., Nissbrandt, H. & Åberg, M. High IQ in Early Adulthood Is Associated with Parkinson ‘s Disease. J. Parkinsons. Dis. 10, 1649–1656 (2020).

28. Domínguez-Baleón, C., Ong, J.-S., Scherzer, C. R., Rentería, M. E. & Dong, X. Understanding the effect of smoking and drinking behavior on Parkinson ‘s disease risk: a Mendelian randomization study. Sci. Rep. 11, 13980 (2021).

29. Noyce, A. J. et al. The Parkinson ‘s Disease Mendelian Randomization Research Portal. Mov. Disord. 34, 1864–1872 (2019).

30. Grover, S. et al. Risky behaviors and Parkinson disease: A mendelian randomization study. Neurology 93, e1412–e1424 (2019).

31. Elbaz, A. et al. Risk tables for parkinsonism and Parkinson ‘s disease. J. Clin. Epidemiol. 55, 25–31 (2002).

32. Miller, K. L. et al. Multimodal population brain imaging in the UK Biobank prospective epidemiological study. Nat. Neurosci. 19, 1523–1536 (2016).

33. Choi, S. W. & O‘Reilly, P. F. PRSice-2: Polygenic Risk Score software for biobank-scale data. Gigascience 8, (2019).

34. Desikan, R. S. et al. An automated labeling system for subdividing the human cerebral cortex on MRI scans into gyral based regions of interest. Neuroimage 31, 968–980 (2006).

35. Alfaro-Almagro, F. et al. Image processing and Quality Control for the first 10,000 brain imaging datasets from UK Biobank. Neuroimage 166, 400–424 (2018).

36. Alfaro-Almagro, F. et al. Confound modelling in UK Biobank brain imaging. bioRxiv 2020.03.11.987693 (2020) doi:10.1101/2020.03.11.987693.

37. Neilson, E. et al. Impact of Polygenic Risk for Schizophrenia on Cortical Structure in UK Biobank. Biol. Psychiatry 86, 536–544 (2019).

38. McIntosh, A. R. & Mišić, B. Multivariate statistical analyses for neuroimaging data. Annu. Rev. Psychol. 64, 499–525 (2013).

39. Winkler, A. M. et al. Cortical thickness or grey matter volume? The importance of selecting the phenotype for imaging genetics studies. Neuroimage 53, 1135–1146 (2010).

40. Yeo, B. T. T. et al. The organization of the human cerebral cortex estimated by intrinsic functional connectivity. J. Neurophysiol. 106, 1125–1165 (2011).

41. Tremblay, C. et al. Brain atrophy progression in Parkinson ‘s disease is shaped by connectivity and local vulnerability. Brain Commun 3, fcab269 (2021).

42. Parkinson Progression Marker Initiative. The Parkinson Progression Marker Initiative (PPMI). Prog. Neurobiol. 95, 629–635 (2011).

43. Zeighami, Y. et al. A clinical-anatomical signature of Parkinson ‘s disease identified with partial least squares and magnetic resonance imaging. Neuroimage 190, 69–78 (2019).

44. Markello, R. D. & Misic, B. Comparing spatial null models for brain maps. Neuroimage 236, 118052 (2021).

45. Grasby, K. L. et al. The genetic architecture of the human cerebral cortex. Science 367, (2020).

46. Hawrylycz, M. J. et al. An anatomically comprehensive atlas of the adult human brain transcriptome. Nature 489, 391–399 (2012).

47. Tremblay, C. et al. Sex effects on brain structure in de novo Parkinson ‘s disease: a multimodal neuroimaging study. Brain 143, 3052–3066 (2020).

48. Jubault, T. et al. Patterns of cortical thickness and surface area in early Parkinson ‘s disease. Neuroimage 55, 462–467 (2011).

49. Gerrits, N. J. H. M. et al. Cortical Thickness, Surface Area and Subcortical Volume Differentially Contribute to Cognitive Heterogeneity in Parkinson ‘s Disease. PLoS One 11, e0148852 (2016).

50. Chen, C.-H. et al. Genetic topography of brain morphology. Proc. Natl. Acad. Sci. U. S. A. 110, 17089–17094 (2013).

51. Rakic, P., Ayoub, A. E., Breunig, J. J. & Dominguez, M. H. Decision by division: making cortical maps. Trends Neurosci. 32, 291–301 (2009).

52. Geschwind, D. H. & Rakic, P. Cortical evolution: judge the brain by its cover. Neuron 80, 633–647 (2013).

53. Fjell, A. M. et al. Development and aging of cortical thickness correspond to genetic organization patterns. Proc. Natl. Acad. Sci. U. S. A. 112, 15462–15467 (2015).

54. Seidlitz, J. et al. Transcriptomic and cellular decoding of regional brain vulnerability to neurogenetic disorders. Nat. Commun. 11, 3358 (2020).

55. Freeman, S. H. et al. Preservation of neuronal number despite age-related cortical brain atrophy in elderly subjects without Alzheimer disease. J. Neuropathol. Exp. Neurol. 67, 1205–1212 (2008).

56. Bishop, N. A., Lu, T. & Yankner, B. A. Neural mechanisms of ageing and cognitive decline. Nature 464, 529–535 (2010).

57. Vidal-Pineiro, D. et al. Cellular correlates of cortical thinning throughout the lifespan. Sci. Rep. 10, 21803 (2020).

58. Patel, Y. et al. Virtual histology of multi-modal magnetic resonance imaging of cerebral cortex in young men. Neuroimage 218, 116968 (2020).

59. McCann, H., Cartwright, H. & Halliday, G. M. Neuropathology of α-synuclein propagation and braak hypothesis. Mov. Disord. 31, 152–160 (2016).

60. Pedersen, K. M., Marner, L., Pakkenberg, H. & Pakkenberg, B. No global loss of neocortical neurons in Parkinson ‘s disease: a quantitative stereological study. Mov. Disord. 20, 164–171 (2005).

61. Soukup, S.-F., Vanhauwaert, R. & Verstreken, P. Parkinson ‘s disease: convergence on synaptic homeostasis. EMBO J. 37, (2018).

62. Raunio, A. et al. Lewy-related pathology exhibits two anatomically and genetically distinct progression patterns: a population-based study of Finns aged 85. Acta Neuropathologica vol. 138 771–782 Preprint at https://doi.org/10.1007/s00401-019-02071-3 (2019).

63. Frigerio, R. et al. Education and occupations preceding Parkinson disease: a population-based case-control study. Neurology 65, 1575–1583 (2005).

64. Chen, H., Schernhammer, E., Schwarzschild, M. A. & Ascherio, A. A prospective study of night shift work, sleep duration, and risk of Parkinson ‘s disease. Am. J. Epidemiol. 163, 726–730 (2006).

65. Simunovic, F., Yi, M., Wang, Y., Stephens, R. & Sonntag, K. C. Evidence for gender-specific transcriptional profiles of nigral dopamine neurons in Parkinson disease. PLoS One 5, e8856 (2010).

66. Bourque, M., Dluzen, D. E. & Di Paolo, T. Neuroprotective actions of sex steroids in Parkinson ‘s disease. Front. Neuroendocrinol. 30, 142–157 (2009).

67. Hirohata, M., Ono, K., Morinaga, A., Ikeda, T. & Yamada, M. Anti-aggregation and fibril-destabilizing effects of sex hormones on alpha-synuclein fibrils in vitro. Exp. Neurol. 217, 434–439 (2009).

68. Purcell, S. et al. PLINK: a tool set for whole-genome association and population-based linkage analyses. Am. J. Hum. Genet. 81, 559–575 (2007).

69. Bycroft, C. et al. The UK Biobank resource with deep phenotyping and genomic data. Nature 562, 203–209 (2018).

70. Krishnan, A., Williams, L. J., McIntosh, A. R. & Abdi, H. Partial Least Squares (PLS) methods for neuroimaging: a tutorial and review. Neuroimage 56, 455–475 (2011).

71. Griffa, A., Alemán-Gómez, Y. & Hagmann, P. Structural and functional connectome from 70 young healthy adults. (2019). doi:10.5281/zenodo.2872624.

72. Shafiei, G. et al. Spatial Patterning of Tissue Volume Loss in Schizophrenia Reflects Brain Network Architecture. Biol. Psychiatry 87, 727–735 (2020).

73. Shin, J. et al. Cell-Specific Gene-Expression Profiles and Cortical Thickness in the Human Brain. Cereb. Cortex 28, 3267–3277 (2018).

74. Arnatkeviciute, A., Fulcher, B. D. & Fornito, A. A practical guide to linking brain-wide gene expression and neuroimaging data. Neuroimage 189, 353–367 (2019).

75. Markello, R., Shafiei, G., Zheng, Y.-Q. & Mišić, B. jabagen: A toolbox for the Allen Brain Atlas genetics data. (2020). doi:10.5281/zenodo.3726257.

76. Hansen, J. Y. et al. Mapping gene transcription and neurocognition across human neocortex. Nat Hum Behav (2021) doi:10.1038/s41562-021-01082-z.

77. Eden, E., Navon, R., Steinfeld, I., Lipson, D. & Yakhini, Z. GOrilla: a tool for discovery and visualization of enriched GO terms in ranked gene lists. BMC Bioinformatics 10, 48 (2009).

78. Mi, H., Muruganujan, A., Casagrande, J. T. & Thomas, P. D. Large-scale gene function analysis with the PANTHER classification system. Nat. Protoc. 8, 1551–1566 (2013).

